# Evaluating lipid-lowering drug targets for Parkinson disease prevention with Mendelian randomization

**DOI:** 10.1101/2020.05.04.20090142

**Authors:** Dylan M. Williams, Sara Bandres-Ciga, Karl Heilbron, the 23andMe Research Team, the International Parkinson’s Disease Genomics Consortium, David Hinds, Alastair J Noyce

## Abstract

**Objective:** To examine whether long-term exposure to statins and other lipid-lowering drugs may affect PD risk – either beneficially or adversely – using Mendelian randomization (MR).

**Methods:** MR analyses were based on variants in genes encoding the targets of several approved or emerging drug classes that reduce circulating low-density lipoprotein cholesterol (LDL-C) or triglycerides. Variants were weighted by their associations with differences in circulating LDL-C, triglycerides or apolipoprotein B (ApoB) using data from genome-wide association studies of lipids (N = 14,004 to 295,826). MR models indexing the effects of modulating each drug target on PD risk were then estimated from genetic associations with PD case-control status (N = 37,688 cases and 981,372 controls).

**Results:** Estimates for statin exposure were incompatible with drug use increasing PD risk, but were not precise enough to confirm a protective effect: odds ratio for PD per standard deviation (SD) reduction in low-density lipoprotein cholesterol = 0.83; 95% confidence interval (CI): 0.65, 1.07. Findings for other LDL-lowering targets were also close to the null. Among triglyceride-lowering targets, variants indexing Apolipoprotein-A5 / Apolipoprotein-C3 modulation suggested a protective effect (OR per SD lower triglycerides = 0.84; 95% CI = 0.80, 0.89), whereas others were null.

**Interpretation:** This genetic evidence does not support findings from large observational studies which suggest that statin exposure could alter risk of PD. Our overall pattern of results suggest peripheral lipid transport may not influence PD etiology, but this does not necessarily exclude effects of statins or the modulation of apolipoproteins A5/C3 via other mechanisms.

## Introduction

Epidemiological studies have found inconsistent associations between statin use and Parkinson disease (PD) risk – indicating that exposure might provide neuroprotection,^1–3^ or heighten PD risk.^4, 5^ However, such observational studies are affected by biases which limit causal inference, such as confounding and reverse causation. Assessing the potential of statins or other lipid-lowering agents for PD prevention robustly with experimental control would be challenging and, hence, the use of other study designs is warranted to examine whether exposure to lipid-lowering drugs mitigates or increases PD risk.

Genetic variation can be used to predict the effects of long-term drug exposure on disease risk. Variation in the vicinity of a protein-coding gene can affect protein production or function in a similar way to the therapeutic modulation of the same protein with drugs. Associations of so-called *cis*-acting variants with traits are not prone to conventional confounding or reverse causation due to Mendelian randomization principles (*cis*-MR).^6^ In this study, we used *cis*-MR to examine whether PD risk may be affected by long-term exposure to several drug classes related to treatment of primary or familial hypercholesterolemia.

## Methods

### Study design

We conducted two-sample MR analyses.^7^ In *cis*-MR models, we principally addressed four licensed lipid-lowering drug classes which reduce circulating low-density lipoprotein cholesterol (LDL-C) (table 1). Given previous evidence that lower Apolipoprotein B (ApoB) concentrations may increase PD risk,^8^ and that ApoB is a major transporter of triglycerides as well as LDL-C, we also addressed a selection of other drug targets that reduce circulating triglycerides and are targeted by licensed or novel therapeutics. For context, we conducted conventional MR models to estimate whether PD risk is affected by long-term reductions in circulating LDL-C, triglycerides and Apolipoprotein-B (ApoB; the lipoprotein involved in LDL-C and triglyceride transport), i.e. estimating expected consequences for PD risk from reductions in these traits irrespective of the means of reduction, and not necessarily due to any specific drug class.

**Table 1.** – Information on lipid-lowering drug targets under investigation.

| Target groups by primary pharmacological action | Examples of drugs /class | Target | Gene encoding target <sup>1</sup> | Gene ID <sup>1</sup> | Gene location |  |  |
| --- | --- | --- | --- | --- | --- | --- | --- |
|  |  |  |  |  | Chromosome | Start coordinate <sup>2</sup> | Stop coordinate <sup>2</sup> |
| Reduced circulating LDL-C | Statins | HMG-coA reductase (HMGCR) | *HMGCR* | 5006 | 5 | 74,632,154 | 74,657,929 |
|  | Evolocumab | Proprotein convertase subtilisin/kexin type 9 (PCSK9) | *PCSK9* | 20001 | 1 | 55,505,221 | 55,530,525 |
|  | Mipomersen | Apo-B 100 messenger RNA (ApoB-100) | *APOB* | 603 | 2 | 21,224,301 | 21,266,945 |
|  | Ezetimibe | Nieman Pick C1-like protein 1 (NPC1L1) | *NPC1L1* | 7897 | 7 | 44,552,134 | 44,580,914 |
| Reduced VLDL-C / triglycerides | Fibrates | Peroxisome proliferator-activated receptor alpha (PPARα) | *PPARA* | 9232 | 22 | 46,546,424 | 46,639,653 |
|  | Evinacumab (investigational) | Angiopoietin-related protein 3 (ANGPTL3) | *ANGPTL3* | 491 | 1 | 63,063,158 | 63,071,830 |
|  | Investigational target | Lipoprotein lipase (LPL) | *LPL* | 6677 | 8 | 19,759,228 | 19,824,770 |
|  | Investigational target | Apolipoprotein A-5 (ApoA5) <sup>3</sup> | *APOA5* | 17288 | 11 | 116,660,083 | 116,663,136 |
|  | Investigational target | Apolipoprotein C-3 (ApoC3) <sup>3</sup> | *APOC3* | 610 | 11 | 116,700,422 | 116,703,788 |

<sup>1</sup> Gene symbols and IDs from the HUGO Gene Nomenclature Committee
<sup>2</sup> Coordinates as per Human Genome reference release GRCh37 / hg19 in UCSC annotation

### SNP-lipid associations

To validate that genetic variants index physiological responses expected from the use of corresponding drugs, we combined genetic association statistics from specific gene regions of interest from three large genome-wide association studies (GWAS) of LDL-C, triglycerides and ApoB (N per SNP-lipid association = 14,004 to 295,826; described in the supporting information).^9–11^

### SNP-PD associations

SNP-lipid association statistics were harmonized with corresponding SNP-PD risk estimates from several large PD case-control GWAS samples using data from 23andMe and the International Parkinson’s Disease Genomics Consortium (total N= 37,688 cases and 981,372 controls; see supporting information, supplemental table 1).^12^ PD ascertainment was via self-report in the 23andMe sample and via clinical assessment in all other samples. Lipid and PD GWAS sample overlap is discussed in the supporting information.

### Statistics

We conducted two types of *cis*-MR models – primarily using adjusted estimates from correlated variants within gene regions, and with secondary analyses using only uncorrelated variants to check consistency (see supporting information for SNP selection and model details).

Effects of general, long-term reductions in LDL-C, triglycerides and ApoB on PD risk were estimated with conventional two-sample MR methods: i) inverse variance weighting (IVW) primarily, and ii) weighted median and MR Egger methods in secondary analyses.^13, 14^

All results were expressed as PD risk per standard deviation (SD) lower LDL-C, triglycerides, or ApoB, so that findings are indicative of the use of corresponding lipid-lowering therapeutics. Power calculations are described in the supporting information. Analyses were conducted in R with packages *‘TwoSampleMR*’ and *‘MendelianRandomization*’.^15, 16^

### Ethics

This study used existing summary GWAS data, so separate ethical approval was not required (all prior studies had ethical approval in accordance with the Declaration of Helsinki).

## Results

A reduction in LDL-C (achievable through any means) was not estimated to affect PD risk (figure 1). Results from alternate methods were consistently null (supplemental table 2). In specific drug target models (figure 1), the point estimate for HMGCR inhibition (indicative of statin use) was consistent with modest neuroprotection, but the confidence interval was wide and included the null. Results for PCSK9 and NPC1L1 inhibition were close to the null. The estimate for ApoB silencing (via mipomersen exposure, for example) was on the side of harm but its confidence interval also included the null. Gene region findings were similar, but less precise, when repeated with LD clumping and IVW to account for SNP correlations (supplemental table 3).

**Figure 1.**
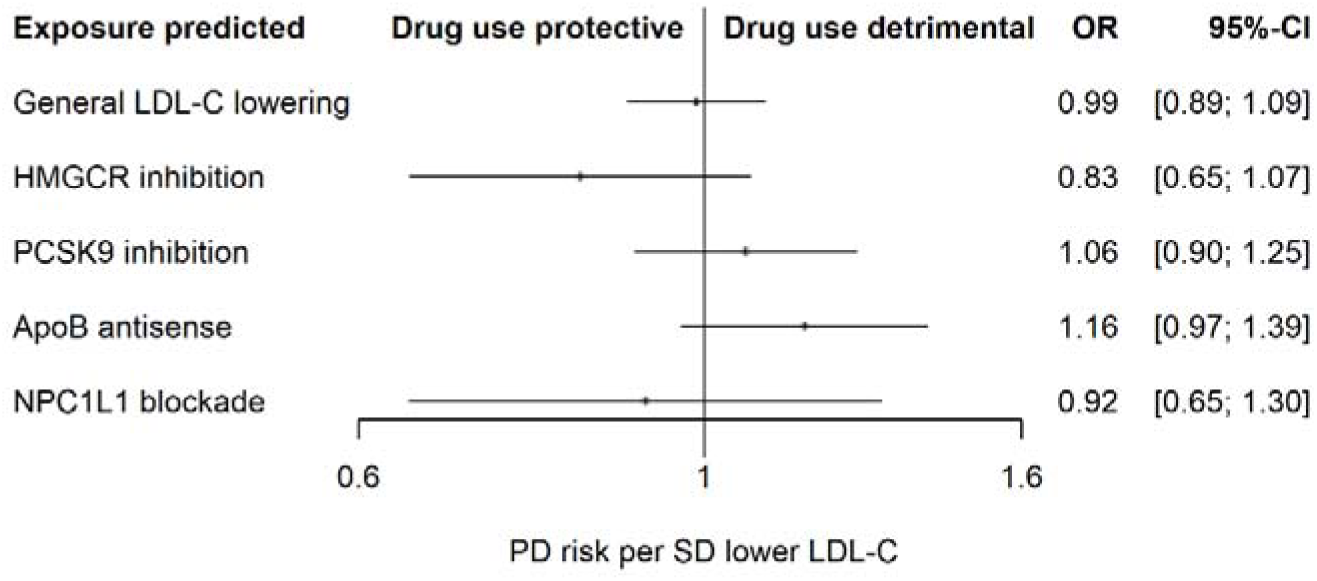
– MR estimates for the effects of a general reduction in LDL-C, and the modulation of LDL-lowering drug targets, on PD risk, weighted by circulating LDL-C.

A reduction in circulating triglycerides was not predicted to affect PD risk, with a point estimate narrowly on the side of harm but close to the null in all models (odds ratio per SD lower circulating triglycerides = 1.06 [95% confidence interval: 0.96, 1.16]; supplemental figure 1 and supplemental table 3). For triglyceride-reducing drug targets (supplemental figure 1), findings were centred on, or close to, the null for *APOB* (ApoB silencing indexed here via a reduction in triglycerides, rather than LDL-C), *PPARA* (indicative of fibrate use), *ANGPTL3* (indicative of angiopoietin-like 3 inhibitors, an investigational class) and *LPL* (indicative of investigational lipoprotein lipase activators). However, many of these results were estimated imprecisely. In contrast, ApoA5 /ApoC3 modulation was predicted to lower PD risk, and estimated with higher precision (odds ratio per SD lower triglycerides from ApoA5/ApoC3 modulation = 0.82 [0.72, 0.94], *P*=0.005).

Finally, models were repeated with genetic indexing of the degree to which all targets under investigation lower ApoB in circulation, reflecting combined reductions in LDL-C and very low-density lipoprotein cholesterol, a major transporter of triglycerides (figure 2 and supplemental table 3). A reduction in circulating ApoB was not estimated to affect PD risk by any MR method, and specific inhibition of ApoB function (averaged across any/all tissue-specific effects) was not notably different. Results for all other LDL- and triglyceride-lowering targets were again close to the null, with the exception of ApoA5 / ApoC3 modulation. The lowest *P* value for genotype-PD associations in the region was 5.9×10^−5^ for rs4520 in *APOC3*.

**Figure 2.**
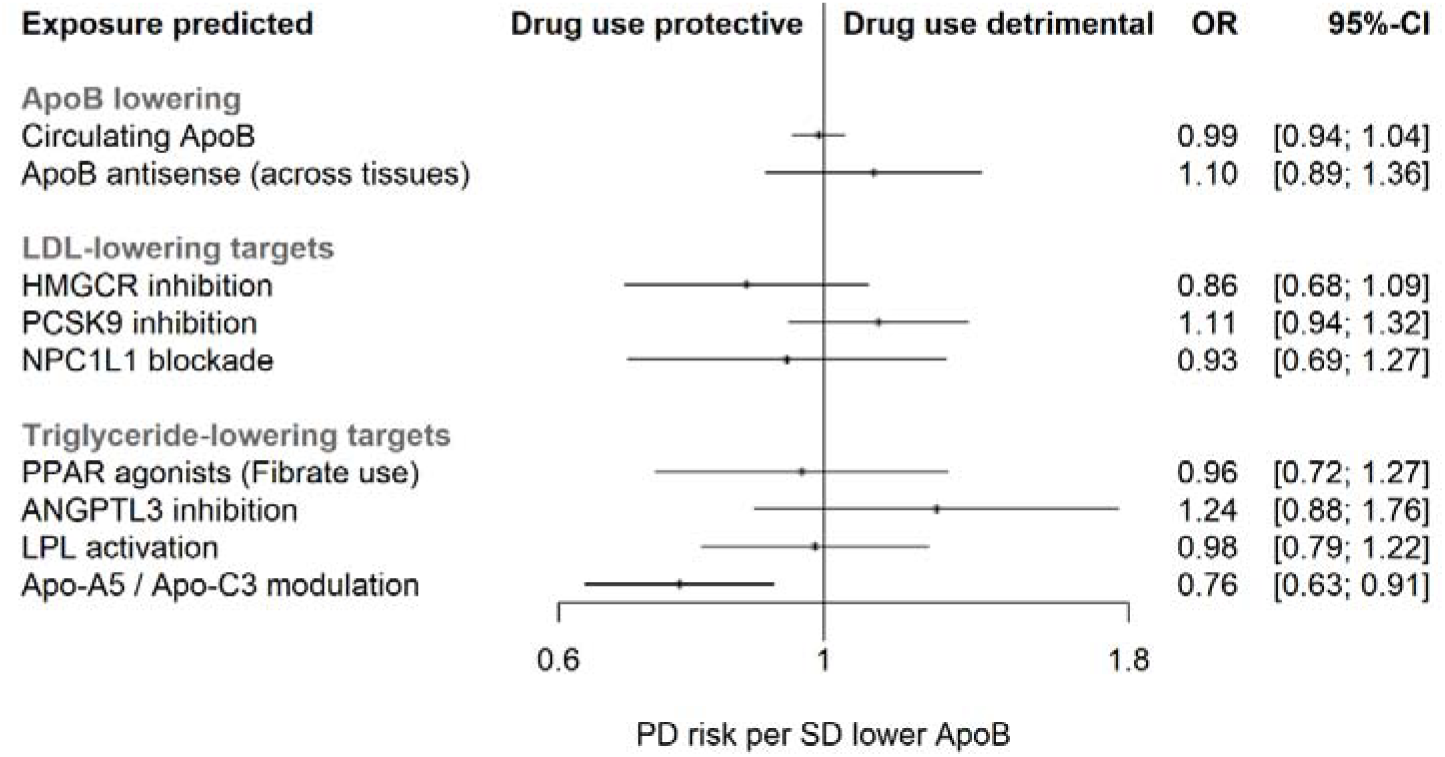
– MR estimates for the effects of modulating lipid-lowering drug targets on PD risk, weighted by circulating ApoB.

Most MR models were expected to have ≥ 80% power to detect odds ratios of 0.88 or lower per SD lipid reduction (supporting information / supplemental table 4).

## Discussion

This is the largest MR study to have investigated lipid-lowering and the modulation of corresponding drug targets in relation to PD risk. Null findings for circulating concentrations of LDL-C, triglycerides and ApoB refute a role of peripheral lipid modulation in the etiology of PD, providing a timely context to conflicting evidence from case-control or prospective studies of lipid biomarkers or statin use.^1–4, 8^ Two previous studies have examined similar questions with MR.^8, 17^ Benn *et al*. did not find effects of lowering LDL-C and modulating HMGCR and PCSK9 on PD risk, but their analyses lacked power to detect effects given MR models based on ≤ 579 PD cases.^17^ Fang *et al*. reported very modest associations of higher genetically-indexed LDL-C (OR = 0.96 [95% CI: 0.92, 0.99]) and triglycerides (OR = 0.94 [95% CI: 0.89, 1.00]) with lower PD risk in a similar sample used for this analysis.^8^ However, these results may have been false positives for technical reasons (too many variants were included in MR models because some are correlated). Even if accurate, the reported magnitudes of effects may imply limited clinical relevance.

Results for specific targets could still indicate benefits (or risks) for PD of using related therapeutics, as their modulation may have physiological effects beyond lipid lowering. Hence, our result for *HMGCR*, on the side of neuroprotection, does not rule out possible benefits of on-target effects of using statins for PD prevention via alternative mechanisms. Concomitantly, this evidence opposes findings from large observational studies of health insurance records that have suggested statin exposure may increase PD risk.^4, 5^ We also identified strong genetic evidence implicating one or more genes in the apolipoprotein gene cluster on chromosome 11 in PD etiology. However, closely situated variants could have *cis*-acting effects on multiple proteins in the region (horizontal pleiotropy) in addition to ApoA5 and ApoC3, and/or be correlated with variants affecting other proteins via linkage. Thus, the current analyses do not pinpoint the exact protein(s) of relevance for PD among several possible candidates encoded from genes in this region.

The major strengths of this study are the use of a very large sample of genetic data and an MR design for causal inference. Key limitations include: i) the estimation of on-target effects of drug use only (these MR models do not estimate potential off-target effects for classes); ii) lack of precision to robustly identify modest but potentially meaningful effects for some targets, including HMGCR inhibition; iii) lifelong, averaged estimates will not identify any critical periods of exposure that targets may have on PD risk. We also note that our findings relate primarily to PD incidence, and not to the potential for these drug targets to mitigate PD progression among cases.

Larger-scale MR studies, and other forms of pharmacoepidemiology, would help to further evaluate the role of statin exposure in PD risk and progression and assess the potential neuroprotective ApoA5 and/or ApoC3 modulation. In particular, using tissue-specific expression quantitative trait loci in future *cis*-MR analyses may help to resolve questions regarding apolipoproteins coded by the *APOA5/APOC3* cluster in relation to the pathogenesis of PD.

## Data Availability

The genetic association summary statistics for PD risk used in these analyses are available via applications to 23andMe and the IPDGC

## Abbreviations

ApoA5: Apolipoprotein A5
ApoC3: Apolipoprotein C3
ApoB / *APOB*: apolipoprotein B
GWAS: genome-wide association study
HMGCR: HMG-CoA reductase
IVW: inverse variance weighted method
LDL-C: low-density lipoprotein cholesterol
MR: Mendelian randomization
PD: Parkinson disease
SNP: single nucleotide polymorphism

## Acknowledgment

DMW received support for this research from the Swedish Research Council (Vetenskapsrådet; grant number 2017-02175), and is funded by the UK’s Medical Research Council (MC_UU_00019/2). AJN is funded by the Barts Charity. We thank the participants from all cohorts who contributed to this study. This research was conducted using data from the IPDGC and 23andMe. Members of the IPDGC and the 23andMe Research Team are listed in the supporting information.

## Author Contributions

DMW, SBC and AJN conceived of the study. All authors contributed to its design. DMW, KH, DH, the IPDGC and 23andMe Research Team contributed to data analyses. All authors contributed to the drafting of the article’s text and approved the manuscript for publication.

## References

1. Undela K, Gudala K, Malla S, Bansal D. Statin use and risk of Parkinson’s disease: a meta-analysis of observational studies. J Neurol. 2013;260(1):158–65.

2. Sheng Z, Jia X, Kang M. Statin use and risk of Parkinson’s disease: A meta-analysis. Behav Brain Res. 2016;309:29–34.

3. Yan J, Qiao L, Tian J, et al. Effect of statins on Parkinson’s disease: A systematic review and meta-analysis. Medicine (Baltimore). 2019;98(12):e14852.

4. Jeong S-M, Jang W, Shin DW. Association of statin use with Parkinson’s disease: Dose-response relationship. Mov Disord. 2019;34(7):1014–21.

5. Liu G, Sterling NW, Kong L, et al. Statins may facilitate Parkinson’s disease: Insight gained from a large, national claims database. Mov Disord. 2017;32(6):913–7.

6. Swerdlow DI, Kuchenbaecker KB, Shah S, et al. Selecting instruments for Mendelian randomization in the wake of genome-wide association studies. Int J Epidemiol. 2016;45(5):1600–16.

7. Burgess S, Butterworth A, Thompson SG. Mendelian randomization analysis with multiple genetic variants using summarized data. Genet Epidemiol. 2013;37(7):658–65.

8. Fang F, Zhan Y, Hammar N, et al. Lipids, Apolipoproteins, and the Risk of Parkinson Disease: A Prospective Cohort Study and a Mendelian Randomization Analysis. Circ Res. 2019;125(6):643–52.

9. Willer CJ, Schmidt EM, Sengupta S, et al. Discovery and refinement of loci associated with lipid levels. Nat Genet. 2013;45(11):1274.

10. Kettunen J, Demirkan A, Würtz P, et al. Genome-wide study for circulating metabolites identifies 62 loci and reveals novel systemic effects of LPA. Nat Commun. 2016;7:11122.

11. Liu DJ, Peloso GM, Yu H, et al. Exome-wide association study of plasma lipids in> 300,000 individuals. Nat Genet. 2017;49(12):1758.

12. Nalls MA, Blauwendraat C, Vallerga CL, et al. Identification of novel risk loci, causal insights, and heritable risk for Parkinson’s disease: a meta-analysis of genome-wide association studies. Lancet Neurol. 2019;18(12):1091–102.

13. Bowden J, Davey Smith G, Burgess S. Mendelian randomization with invalid instruments: effect estimation and bias detection through Egger regression. Int J Epidemiol. 2015;44(2):512–25.

14. Bowden J, Davey Smith G, Haycock PC, Burgess S. Consistent estimation in Mendelian randomization with some invalid instruments using a weighted median estimator. Genet Epidemiol. 2016;40(4):304–14.

15. Hemani G, Zheng J, Elsworth B, et al. The MR-Base platform supports systematic causal inference across the human phenome. eLife. 2018;7:e34408.

16. Yavorska OO, Burgess S. MendelianRandomization: an R package for performing Mendelian randomization analyses using summarized data. Int J Epidemiol. 2017:dyx034.

17. Benn M, Nordestgaard BG, Frikke-Schmidt R, Tybjærg-Hansen A. Low LDL cholesterol, PCSK9 and HMGCR genetic variation, and risk of Alzheimer’s disease and Parkinson’s disease: Mendelian randomisation study. BMJ. 2017;357:j1648.

